# Wastewater-based sequencing of Respiratory Syncytial Virus enables tracking of lineages and identifying mutations at antigenic sites

**DOI:** 10.1101/2025.02.28.25321637

**Authors:** Jolinda de Korne-Elenbaas, Auguste Rimaite, Ivan Topolsky, David Dreifuss, Charlyne Bürki, Lara Fuhrmann, Louis du Plessis, William J. Fitzsimmons, Emily E. Bendall, Tanja Stadler, Niko Beerenwinkel, Timothy R Julian

## Abstract

Respiratory Syncytial Virus (RSV) infections pose a substantial health burden, especially for vulnerable populations such as infants and the elderly. While novel immunoprophylaxis offer promising protection, many countries lack robust surveillance systems to monitor circulating RSV lineages and detect mutations that might compromise the effectiveness of these new interventions. In this study, we applied an RSV subtype-specific amplicon-based sequencing approach to obtain RSV-A and RSV-B sequences from wastewater, providing catchment-wide insights into circulating RSV lineages. RSV was amplified and sequenced from wastewater samples collected during the 2022-2023 and 2023-2024 RSV seasons from wastewater treatment plants in Zurich and Geneva, Switzerland. During the 2022-2023 season, RSV-B was the predominant subtype, with the B.D.E.1 lineage prevailing in both cities. In the 2023-2024 season, RSV-A predominated and wastewater sequence data revealed co-circulation of multiple lineages, including A.D.1, A.D.3, A.D.5, and their sub-lineages. Wastewater-derived fusion gene sequences revealed several amino acid substitutions compared to the reference genomes. Most of these corresponded to known signatures of circulating lineages. However, low-frequency, non-synonymous mutations in antigenic sites of both RSV-A and RSV-B were also identified, of which some had not been reported in clinical sequences. These findings demonstrate the potential of wastewater-based genomic surveillance to identify and track circulating RSV lineages and clinically relevant mutations. As novel RSV immunoprophylaxis measures are introduced in upcoming RSV seasons, wastewater-derived genomic RSV data provides a valuable baseline for understanding RSV diversity and future viral evolution under immunological pressure.

## Introduction

Respiratory Syncytial Virus (RSV) is a common respiratory pathogen that typically causes mild, cold-like symptoms. However, in vulnerable populations such as infants and older adults, RSV infections can progress to severe respiratory disease. These severe RSV infections may result in hospitalization and even death^1,2^, placing a substantial burden on healthcare systems worldwide^3,4^. The prevention of severe respiratory disease caused by RSV has traditionally relied on a passive immunization strategy designed to protect infants with underlying health conditions^5^. Recently, novel RSV vaccines and monoclonal antibodies have demonstrated efficacy in reducing hospitalizations due to RSV-associated lower respiratory tract infections, leading to their approval in multiple countries^6–8^.

Current monitoring of RSV infections often relies on voluntary case reporting by clinicians^9^, with routine RSV testing primarily focused on vulnerable populations in hospitals and elderly care facilities. Therefore, existing surveillance systems are incomplete and biased towards severe clinical cases, resulting in a limited understanding of the virus’ prevalence in the general population. In several countries, clinical RSV surveillance systems have recently been complemented by the monitoring of RSV RNA concentrations in wastewater^10–14^. Wastewater-based surveillance has been shown to be effective for tracking pathogens with a large public health impact, as it provides unique, population-level insights into pathogen prevalence and spread^15–19^. Studies have demonstrated that RSV concentrations in wastewater closely mirror trends in reported clinical cases, supporting its reliability as a surveillance method^11,20^. Especially in regions with limited clinical surveillance, wastewater-based monitoring could offer a resource for tracking RSV seasonality and informing the timing of immunization programs^21^.

The introduction of novel RSV vaccines underscores the importance of genomic RSV surveillance, in addition to monitoring RNA concentrations, for tracking circulating strains and potential vaccine resistance mutations. RSV is classified into two major antigenic groups, RSV-A and RSV-B, which typically alternate predominance every one to two seasons^22,23^. Besides subtyping, clinical studies have implemented RSV full-genome sequencing to investigate molecular epidemiology of RSV and track its evolution^24,25^. However, genomic sequencing of RSV remains rare. Upon the start of this study, only a single full-genome sequence of RSV-B from Switzerland was publicly available, dating from 2019^26^.

Wastewater-based approaches have the potential to provide population-level genomic surveillance, as demonstrated for SARS-CoV-2^27^. Studies have shown the ability to subtype RSV and sequence part of its surface glycoprotein (G) gene directly from wastewater^14,28^. Here, we extended these efforts and developed an approach for whole-genome sequencing of RSV from wastewater, using an amplicon-based sequencing method. We demonstrate sufficient coverage to track the relative abundance of distinct RSV-A and RSV-B lineages and monitor the antigenic sites targeted by the vaccines. This strategy enhances our understanding of RSV molecular epidemiology in large populations, providing insights into circulating strains prior to the introduction of novel RSV vaccines.

## Methods

### Wastewater sampling and processing

Twenty four-hour composite raw influent wastewater samples were collected from wastewater treatment plants (WWTPs) in Zurich (serving 471,000 people) and Geneva (serving 454,000 people), as part of the Swiss wastewater-based respiratory virus monitoring program (Figure 1)^11^. Total nucleic acids were extracted and concentrated from 40mL influent wastewater into an elution volume of 80μL with a vacuum-based direct capture method using the Wizard® Enviro Total Nucleic Acid Extraction kit (Promega Corporation, Madison, USA). Extracts were further purified using OneStep PCR Inhibitor Removal columns (Zymo Research, Irvine, USA). Extracts were diluted three times to further reduce PCR inhibitor concentrations and stored at -80°C for up to 1.5 years before sequencing.

**Figure 1.**
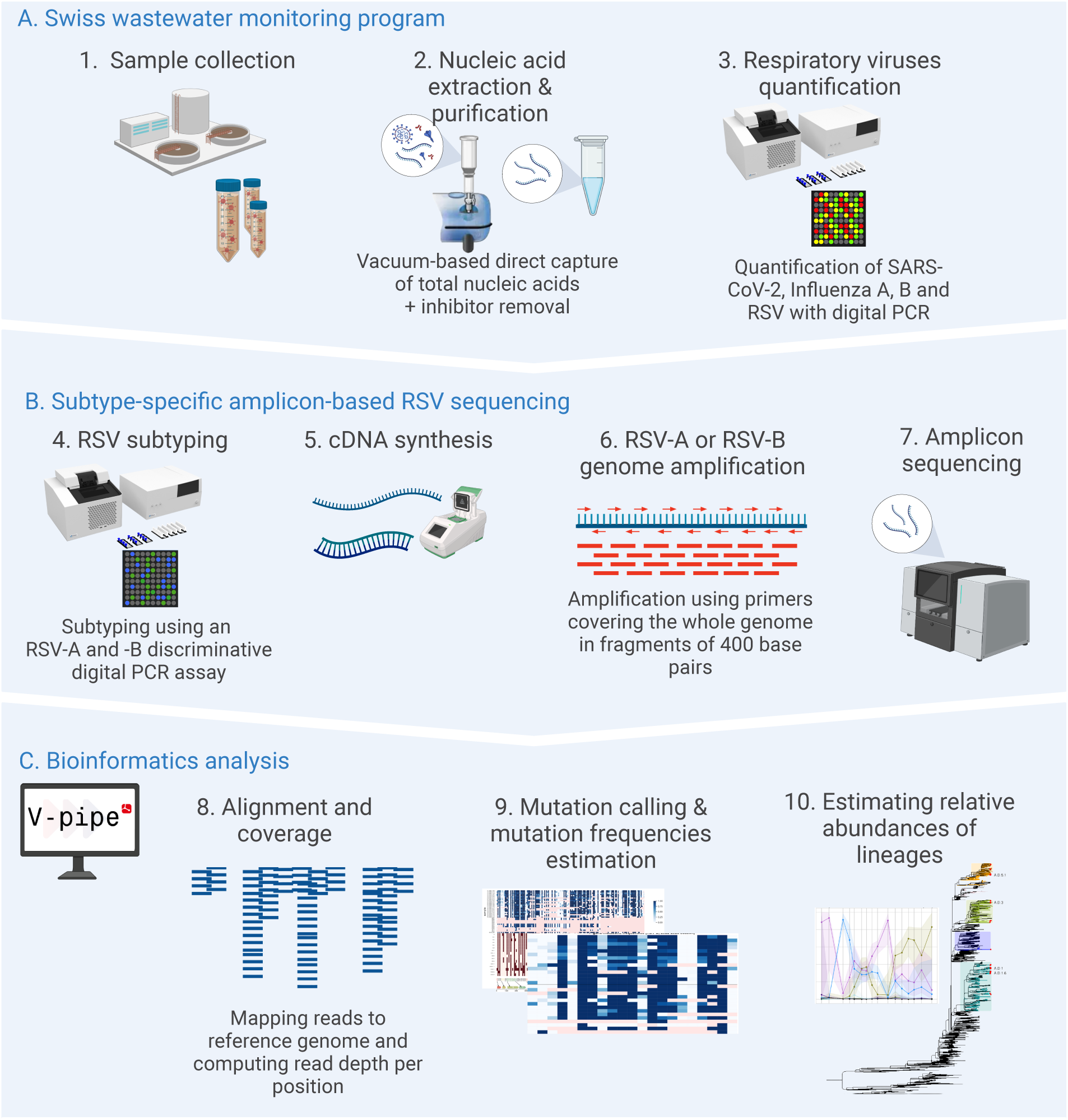
Workflow for amplicon-based subtype-specific RSV whole genome sequencing from wastewater. The workflow consists of three parts: (A) wastewater sample processing as part of the Swiss national wastewater monitoring program for respiratory viruses, (B) the wet-lab process for wastewater-based RSV sequencing, and (C) bioinformatics analysis of wastewater-derived RSV sequence data using the V-pipe computational pipeline.

### Sample selection

RSV was quantified with a pan-RSV digital PCR (dPCR) assay in wastewater extracts from September 2022 through April 2024. Seasonal waves in RSV concentrations in wastewater were observed in Zurich and Geneva from October 2022 until April 2023, and again from November 2023 until April 2024 (Figure S1). During these periods, one extract per location per week was selected for sequencing, prioritizing the extract with the highest or second-highest measured RSV concentration. In total, 14 samples were selected from both Zurich and Geneva for the 2022-2023 season and 18 samples per location for the 2023-2024 season (Figure S1).

### RSV subtyping

To select the set of amplification primers to use (RSV-A or RSV-B), the dominant RSV subtype of the season was determined in a subset of samples using the assay described by Wang et al^29^. The RSV-A probe was modified to improve the signal for dPCR (Table S1). The assay was prepared using a pre-reaction volume of 21.6μL mastermix (Table S2) and 5.4μL diluted extract, of which a reaction volume of 25μL was loaded onto Sapphire chips (Stilla Technologies, France). The assay was run on the Naica® system for Crystal Digital PCR (Stilla Technologies, France), using a Geode for partitioning and thermocycling, and the Prism3 for fluorescent imaging. Within each Geode, a positive control and a no template control (NTC) were run alongside up to ten samples. A sample passed quality control if it had more than 15,000 analyzable droplets, the positive control contained droplets positive for both subtypes, and the NTC had fewer than 3 positive droplets for both subtypes.

### Design of amplicon-based RSV sequencing method

Starting from an RSV subtype-specific amplicon-based sequencing method initially established for clinical samples^30^, we designed an optimized protocol for use on wastewater extracts. The optimization included modification of primers to improve low coverage and avoid dropouts of amplicons by taking into account the mutations occurring at the binding sites of these primers on recent European genomes PP970044.1 and OQ261752.1 (RSV-A), and MT107528.1 (RSV-B). Uniformity of coverage depth across the genome for RSV-B was optimized by adjusting individual primer concentrations according to amplicon coverage such that lower concentrations of primers were used for amplicons that were relatively overrepresented and higher concentrations for underrepresented amplicons^31^. The coordinates of the primers and inserts were based on commonly used reference genomes for RSV-A hRSV/A/England/397/2017 (GISAID accession no. EPI_ISL_412866) and RSV-B HRSV/B/AUS/VIC-RCH056/2019 (GISAID accession no. EPI_ISL_1653999).

### RSV amplification and sequencing

The sequencing workflow (Figure 1) begins with reverse transcription of RNA using LunaScript RT SuperMix (New England Biolabs) (Table S3), followed by amplification of the complementary DNA with Q5 Hot Start High-Fidelity 2X Master Mix (New England Biolabs) (Table S4). The full RSV genome was amplified in fragments of 400 bp, using 50 (RSV-A) or 51 (RSV-B) primer sets (Table S5). To prevent overlapping amplicons in one reaction, primer sets were divided into two pools and amplification occurred in two separate reactions for each sample. Amplicons of these two reactions were subsequently pooled and analyzed on a D1000 ScreenTape (Agilent Technologies) to check for the presence of 400 bp fragments. An NTC, in which the extract was replaced with water, was included in each amplification run. The absence of a band in the NTC was used to confirm absence of cross-contamination. The resulting 400 bp amplicons were sequenced on either NextSeq2000 (Illumina) or AVITI (Element Biosciences) sequencing systems, yielding paired-end reads of length 250 bp.

### Alignment and coverage

Paired-end FASTQ files containing raw sequencing data were processed using the viral next-generation sequencing data analysis software V-pipe version 3.0.0.pre1 (Figure 1)^32^. Configuration details are provided in the config.yaml file, and data analysis and visualization scripts (Python versions 3.11.8 and 3.12.6, R version 4.3.2) are available on Github. In short, data preprocessing, including trimming and filtering low-quality reads and trimming adapters, was performed using PRINSEQ^33^ with the default V-pipe settings. Reads were aligned using BWA^34^ against the reference genomes EPI_ISL_412866 for RSV-A and EPI_ISL_1653999 for RSV-B. The reads were classified as RSV-A or RSV-B on the basis of the highest scoring alignment. Reads were filtered out if the length of the aligned region was shorter than 200 base pairs. Primers were trimmed based on their coordinates w.r.t. the reference genomes using the ampliconclip function of SAMtools^35^. Read depths per position of the reference genome were computed from the filtered and trimmed alignment files using the V-pipe built-in function aln2basecnt. For each sample, genome coverage was calculated as the proportion of reference genome positions with read depth more than 30. To assess the relationship between total RNA concentration measured with the pan-RSV dPCR assay (denoted *conc*) and genome coverage (denoted *cov*), we fitted the exponential saturation curve *E*[*cov*] = 1 − *e*^-*conc* * *k*^ with parameter *k* and computed nonparametric Spearman correlation coefficients. The curves were fitted using the robust linear regression − *log* (1 − *cov*) ∼ *conc* by employing the R package robustbase (version 0.99-2).

### Mutation calling and computation of mutation frequencies

Mutations were identified relative to the EPI_ISL_412866 (RSV-A) and EPI_ISL_1653999 (RSV-B) reference genomes using Lofreq version 2.1.3^36^. Mutation frequencies were extracted from the VCF files as the ratio of the read depth for the respective mutation to the total read depth at that position. For visualization purposes, only mutations with frequency above 0.02 in at least two wastewater samples were plotted^37^. Positions with read depth below 30 were considered dropouts and regarded as missing values in further analyses. Nucleotide substitutions were translated into amino acid substitutions using an adapted version of vcf-annotator^38^.

### Signature mutations and relative abundances of lineages

Signature mutations of RSV-A and RSV-B lineages, defined relative to the reference genomes, were identified from publicly available RSV-A and RSV-B consensus genomes available on GenSpectrum^39^ by querying the open LAPIS API^40^. A mutation was considered as a signature mutation for a lineage if it was present in at least 90% of all publicly available RSV clinical sequences (with non-ambiguous nucleotides at that particular position) assigned to that lineage^41^. Signature mutation frequencies in wastewater RSV sequence data were calculated as described above, and deconvolved into the relative abundances of lineages using Lollipop version 0.5.0^42^. Lollipop combines least square optimization with kernel-based smoothing for deconvolution, and is designed to enhance confidence in estimating relative lineage abundances in wastewater by leveraging time series data. The kernel bandwidth parameter was set to 30.0, regressor was set to robust, f_scale was set to 0.01, and min_tol was set to 1e-3. The number of bootstrap samples was set to 100. For RSV-A, the lineages for the deconvolution were predefined from a subset of 51 RSV-A sequences originating from Switzerland between 2023 and 2024, publicly available on the European Nucleotide Archive (ENA) under the project PRJEB83635 (filtered based on whole-genome non-ambiguous nucleotide coverage above 66%). Phylogenetic analysis was performed with an adapted version of the Nextstrain RSV build^43,44^, which includes a Nextclade filter to select only high-quality sequences from all publicly-available RSV-A consensus genomes (n = 2,906)^45^. Based on this subset, the RSV-A lineages included in the deconvolution were A.D.1, A.D.1.5, A.D.1.6, A.D.2.1, A.D.3, A.D.3.1, A.D.5.1, and A.D.5.2 (Figure S2). Since the total number of publicly available Swiss RSV-B sequences was limited (n = 8), all RSV-B lineages that circulated in Europe after G gene duplication^41^ were used for deconvolution, namely B.D, B.D.1, B.D.1.1, B.D.4, B.D.4.1, B.D.4.1.1, B.D.E.1, B.D.E.2, and B.D.E.4. Absolute lineage abundances were determined by scaling lineage proportions to 7-day median flow-normalized total RSV viral loads in wastewater, measured using the same wastewater extracts as part of the Swiss wastewater-based monitoring program for respiratory viruses (data publicly available^46^).

## Results

### RSV subtypes alternated between 2022-2023 and 2023-2024 seasons

Wastewater samples were processed as part of the Swiss wastewater-based respiratory virus monitoring program, prior to the RSV amplicon-based sequencing workflow (Figure 1). Since the amplicon-based RSV sequencing method uses subtype-specific primers, a subset of samples selected for sequencing was subtyped to determine the dominant RSV subtype for each season. For season 2022-2023, all samples from Zurich showed dominance of RSV-B, whereas a subset of samples from Zurich and Geneva of 2023-2024 showed RSV-A as the dominant circulating subtype (Figure S3). Consequently, the dominant subtypes were considered to represent the total RSV concentration of the respective season.

### Genome coverage of RSV-A and RSV-B correlated with pan-RSV concentrations

Based on the results of the RSV subtyping assay, all samples from RSV season 2022-2023 were sequenced for RSV-B. We achieved a median coverage of 88% (range 36-96%) (Figure S4), meaning that 88% of the genomic positions were covered by at least 30 reads. Wastewater samples from the 2023-2024 season were sequenced for RSV-A, which yielded a median coverage of 62% (range 0-91%) (Figure S5). The fusion (F) gene had a median coverage of 96% (range 13-100%) for RSV-B and 62% (range 0-100%) for RSV-A. Full genome coverage was significantly correlated with total RSV concentrations measured in the pan-RSV assay for both RSV-A (Spearman’s ⍴=0.69, p = 2.78×10^-6^, two tailed) and RSV-B (Spearman’s ⍴=0.42, p = 0.027, two tailed) (Figure 2).

**Figure 2.**
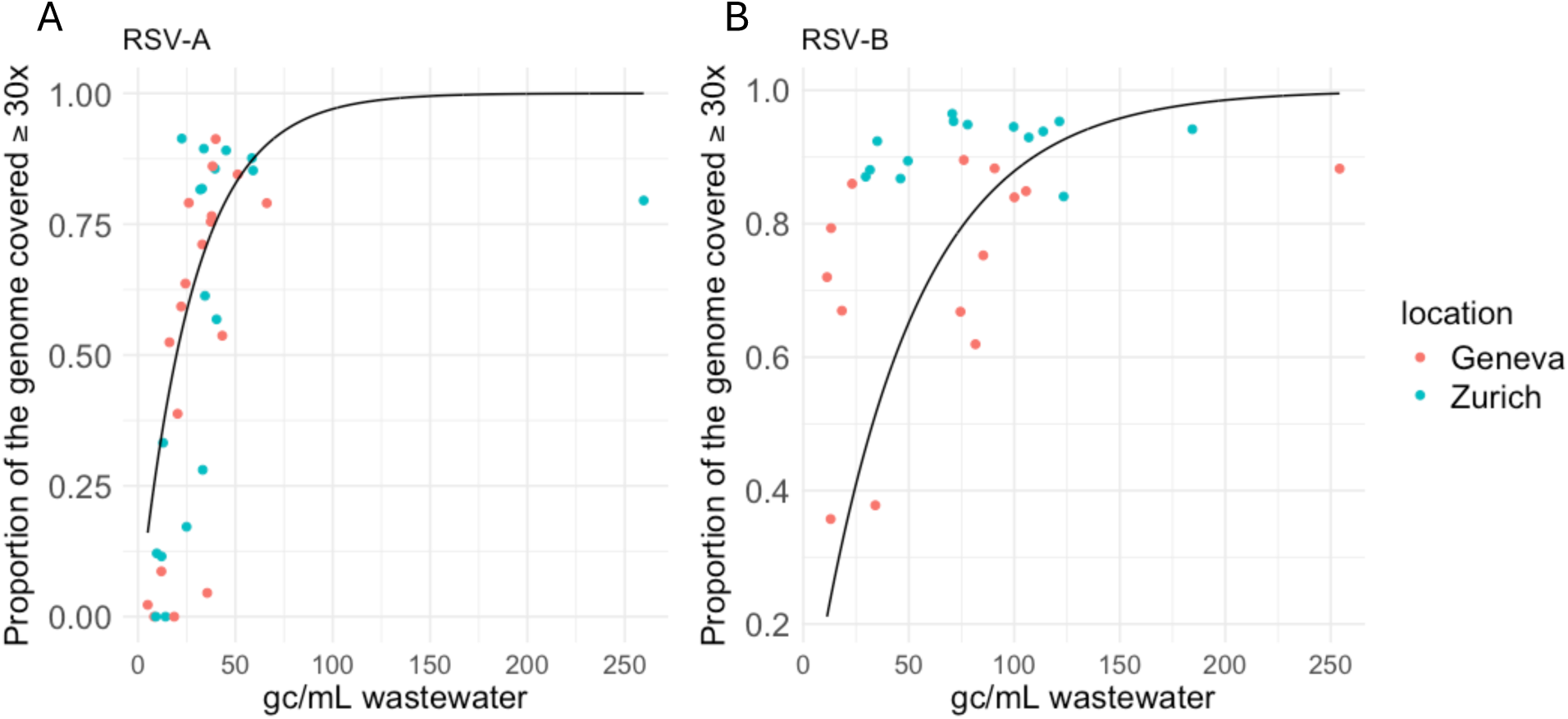
Proportion of the reference genome covered with a read depth of at least 30 for (A) RSV-A and (B) RSV-B versus the total concentrations of RSV, in genome copies per mL wastewater measured with the pan-RSV digital PCR assay. Points correspond to individual samples and colors correspond to the wastewater sampling location. The solid line shows a fitted exponential saturation curve.

### B.D.E.1 was the predominant RSV-B lineage in wastewater samples of the 2022-2023 season

For wastewater samples collected during the 2022-2023 season, the majority of RSV-B mutation frequencies remained consistent over time and across locations. The non-synonymous mutations detected in the samples aligned mainly with the signature mutations of the B.D.E.1 lineage (Figure 3 A,B). When considering all B.D.E.1 signature mutations, both synonymous and non-synonymous, the majority were detected in more than one wastewater sample, with the exception of G1A and C3G, which occur at positions not covered by the amplification primers (Figure S6). Deconvolution of signature mutations into lineages showed a strong predominance of B.D.E.1 during the whole 2022-2023 season in both Zurich and Geneva (Figure 4 A-D).

**Figure 3.**
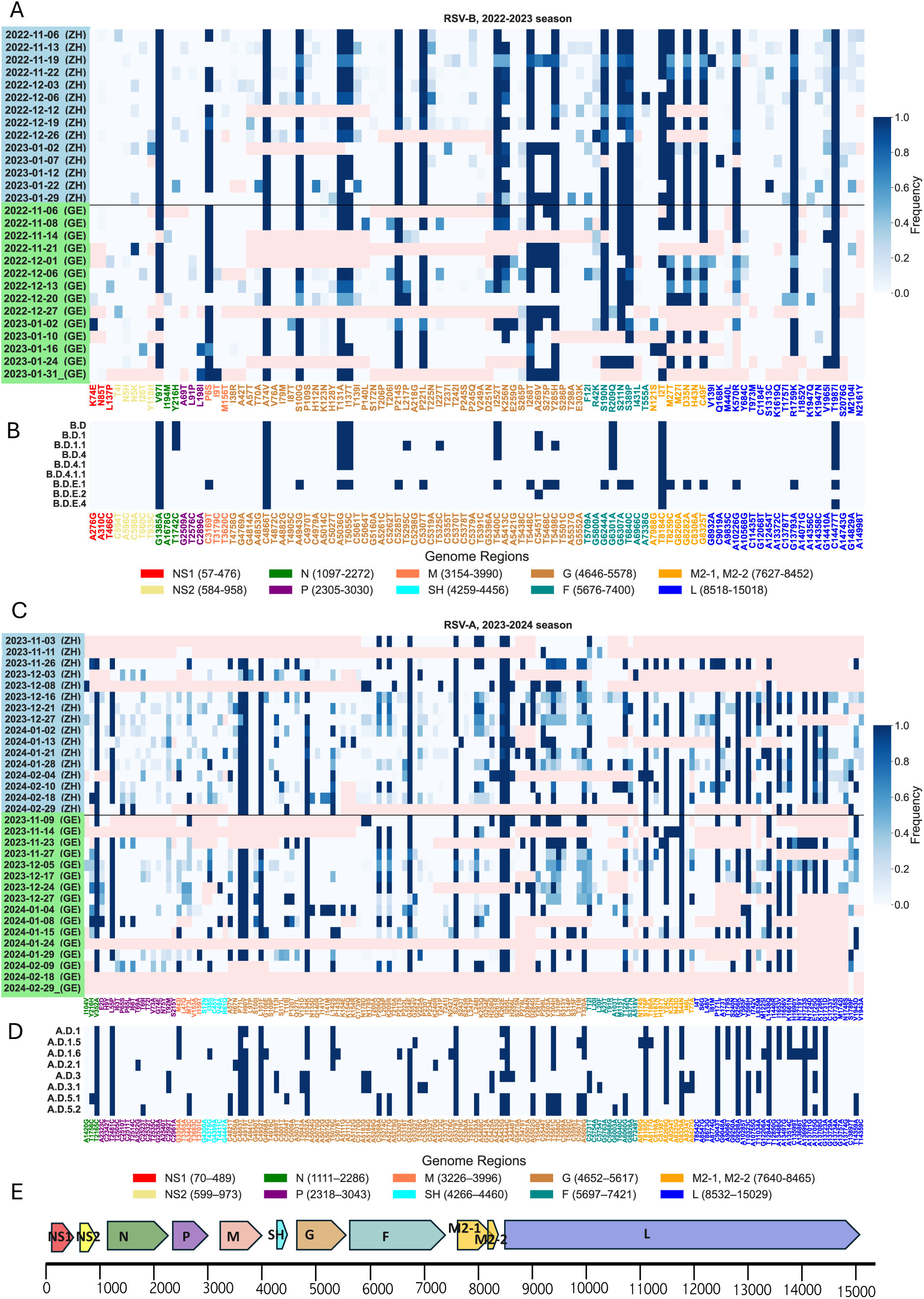
Non-synonymous mutations in wastewater-derived RSV sequences, their frequencies, and their occurrence as signature mutations in RSV lineages. (A) Observed frequencies of non-synonymous mutations in wastewater-derived RSV-B sequences from Zurich (ZH, blue highlighted row labels) and Geneva (GE, green highlighted row labels) obtained during the RSV season of 2022-2023. Rows refer to samples and labels include the date and location of collection. Frequencies are encoded from white (lowest, 0.0) to dark blue (highest, 1.0), while dropouts (read depth below 30 per position) are shown in pink. The visualized mutations appeared in at least two samples with a frequency above 0.02 and a read depth of at least 30. (B) The presence (dark blue) or absence (white) of each non-synonymous mutation as a signature mutation of the RSV-B lineages, which circulated in Europe after G gene duplication. Nucleotide mutations were translated into amino acids and color-coded according to the gene in which they occur. Mutations are reported relative to the reference EPI_ISL_1653999. (C) Observed frequencies of non-synonymous mutations in wastewater-derived RSV-A sequences from Zurich (ZH, blue highlighted row labels) and Geneva (GE, green highlighted row labels) obtained during the RSV season of 2023-2024. (D) The presence (dark blue) or absence (white) of each non-synonymous mutation as a signature mutation of the RSV-A lineages, which were recently detected in Switzerland. (E) Genome structure of RSV with gene positions represented by color-coded arrows.

**Figure 4.**
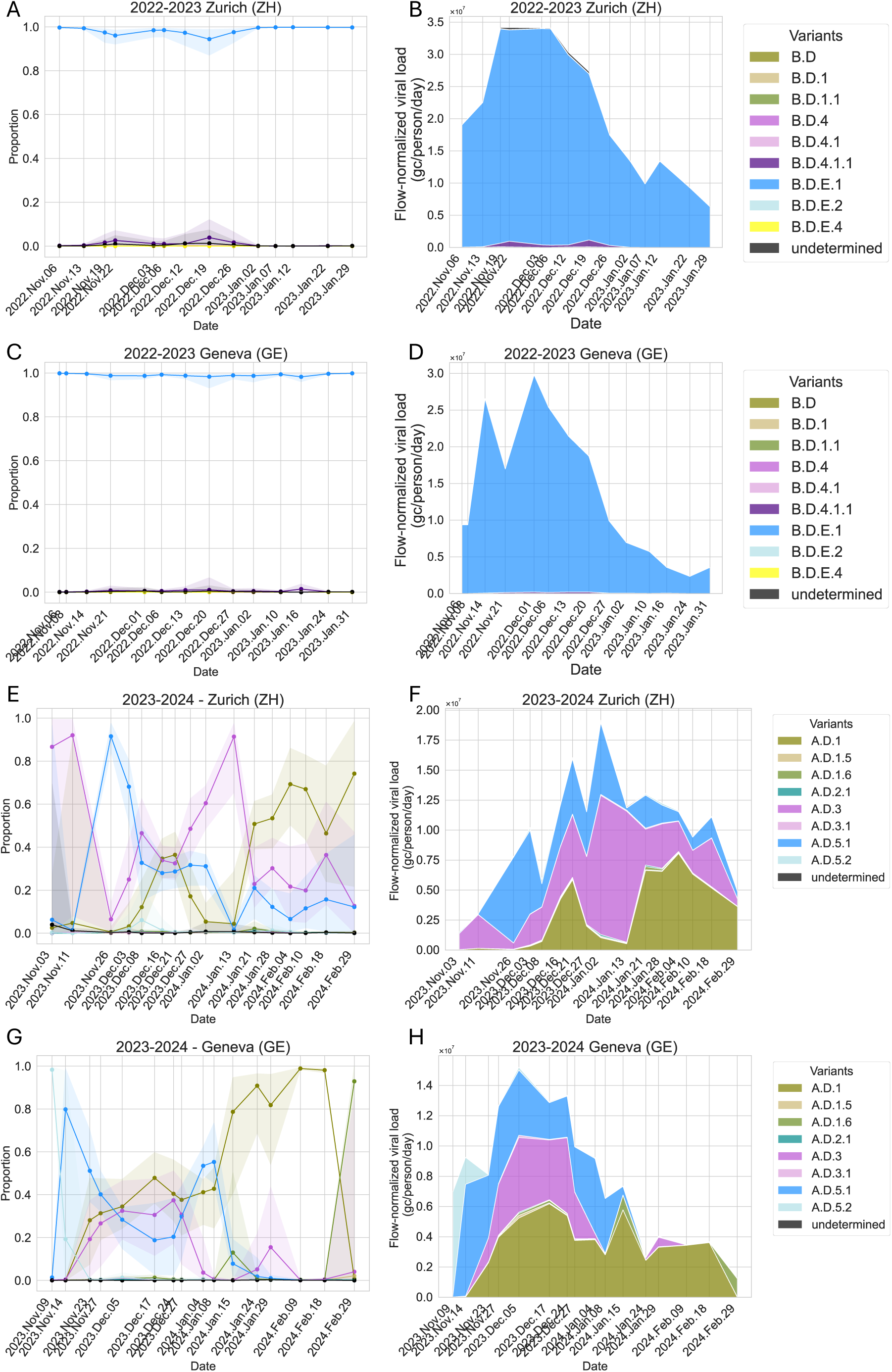
Deconvolution of signature mutation frequencies into lineages enabled tracking temporal dynamics of circulating RSV lineages. (A-D) Estimated proportions and abundances of RSV-B lineages in wastewater collected from Zurich (A: proportions, B: abundances) and Geneva (C: proportions, D: abundances) in the 2022-2023 RSV season. RSV-B lineage B.D.E.1 was predominant during the whole season in both locations. (E-H) Estimated proportions and abundances of RSV-A lineages in wastewater collected from Zurich (E: proportions, F: abundances) and Geneva (G: proportions, H: abundances) in the 2023-2024 RSV season. Multiple RSV-A lineages co-circulated, with A.D.5 sub-lineages mainly circulating early in the season, A.D.1 predominating towards the end of the season, and A.D.3 co-circulating throughout the season in varying relative abundances. The shaded bands around the lines represent the 95% bootstrap confidence intervals around the proportion estimates, based on bootstrapping. Lineage abundances were estimated by scaling lineage proportions to 7-day median flow-normalized total RSV viral loads in wastewater, measured with a pan-RSV assay as part of the Swiss wastewater-based monitoring program. Note that these are approximate viral loads, since co-circulation of RSV-A and RSV-B subtypes is not taken into account.

### Co-circulation of RSV-A lineages in wastewater samples of the 2023-2024 season

Nucleotide substitutions observed in wastewater samples collected during the 2023-2024 season were also consistent over time and across locations, with most observed mutations overlapping with the mutations in recently circulating RSV-A lineages observed in clinical samples in Switzerland (Figure 3 C,D). Deconvolution of signature mutation frequencies into RSV-A lineages detected in Swiss clinical samples showed that multiple RSV-A lineages co-circulated, with the A.D.5 sub-lineages mainly circulating early in the season, A.D.1 predominating towards the end of the season, and A.D.3 co-circulating throughout the season in varying relative abundances (Figure 4 E-H). The patterns of relative abundance of A.D.1 were similar between Zurich and Geneva, with A.D.1 becoming predominant toward the end of the season. However, the abundances of A.D.5 and A.D.3 were not consistent across locations. In Geneva, both lineages were only present in the beginning of the season, whereas in Zurich, they persisted throughout. A.D.3 was already present in Zurich in the first sample and predominated at the beginning of the season, albeit with high uncertainty in the estimate due to low genome coverage (Figure S5).

### RSV-B F gene amino acid substitutions identified in wastewater samples of the 2022-2023 season

Mutations identified in RSV-B F gene sequences from wastewater closely aligned with signature mutations of the B.D.E.1 lineage (Figure 5 A,B). Three amino acid substitutions, S190N, S211N, and S389P, are signature amino acid substitutions for B.D.E.1 and were consistently observed at high frequencies throughout the season in both cities. Low-frequency non-synonymous mutations were also identified in wastewater, resulting in amino acid substitutions F12I, R42K, R209Q, I431L, and T555A. F12I was found in four out of eight publicly available RSV-B sequences from Switzerland (Table S6), all of which are assigned to the B.D.E.1 lineage. R42K and R209Q were absent in recent Swiss clinical sequences but detected in global clinical samples. T555A appeared in only a single global sequence, while I431L was not found in any clinical sequence (Table S6).

**Figure 5.**
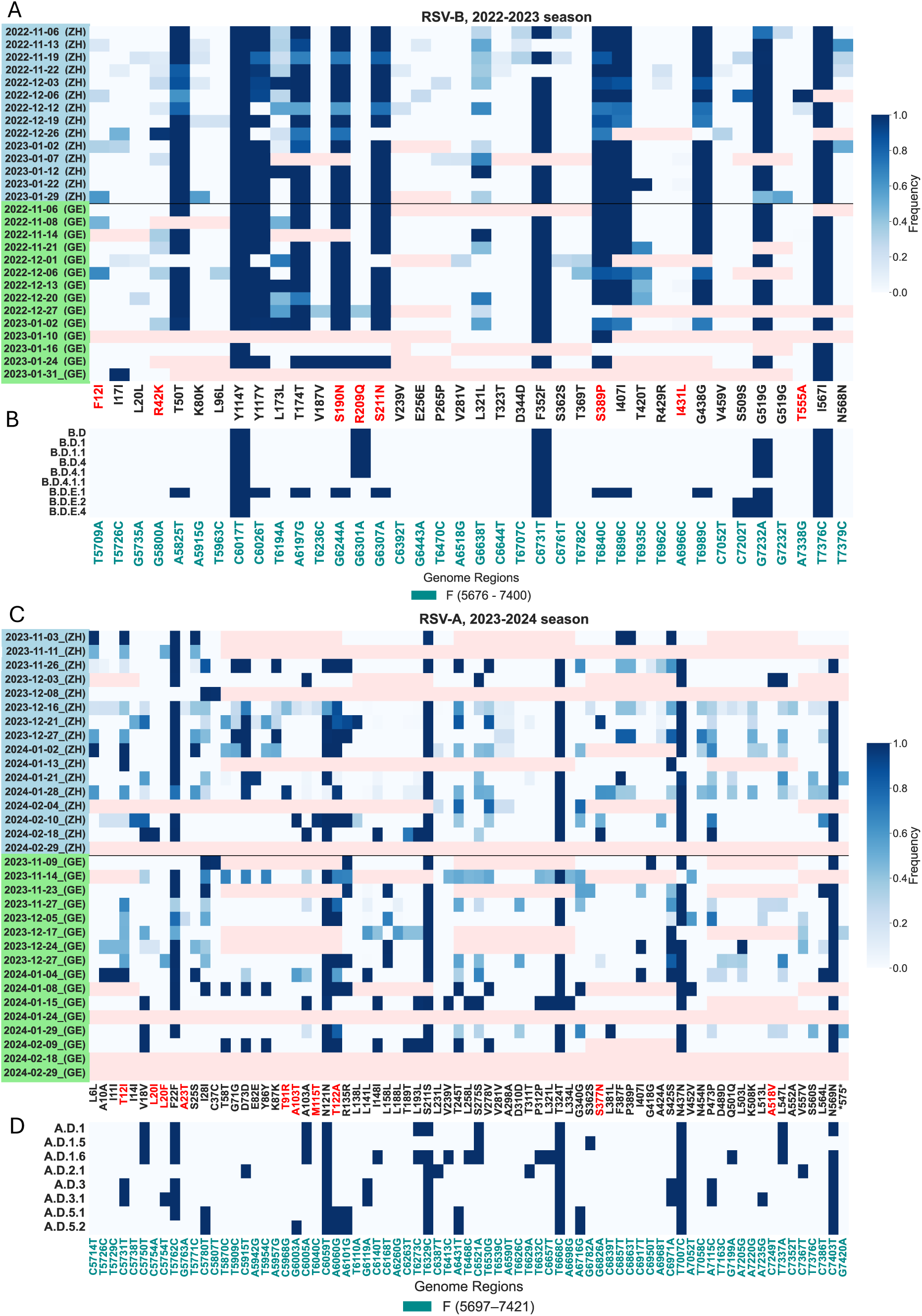
Mutations in wastewater-derived RSV fusion (F) gene sequences, their frequencies, and their occurrence as signature mutations in RSV lineages. (A) Observed frequencies of mutations (both synonymous and non-synonymous) in wastewater-derived RSV-B F gene sequences from Zurich (ZH, blue highlighted row labels) and Geneva (GE, green highlighted row labels) obtained in the 2022-2023 RSV season. Rows refer to samples and labels include the date and location of collection. Frequencies are encoded from white (lowest, 0.0) to dark blue (highest, 1.0), while dropouts (read depth below 30 per position) are shown in pink. The visualized mutations appeared in at least two samples with a frequency above 0.02 and a read depth of at least 30. (B) The presence (dark blue) or absence (white) of each mutation as a signature mutation of the RSV-B lineages, which circulated in Europe after G gene duplication. Nucleotide mutations were translated into amino acids and those in gene-coding regions are color-coded according to the gene in which they occur. Amino acid substitutions are highlighted in red. Mutations are reported relative to the reference EPI_ISL_1653999. (C) Observed frequencies of mutations (both synonymous and non-synonymous) in wastewater-derived RSV-A F gene sequences from Zurich (ZH, blue highlighted row labels) and Geneva (GE, green highlighted row labels) obtained in the RSV season 2023-2024. (D) The presence (dark blue) or absence (white) as a signature mutation of the RSV-A lineages, which were recently detected in Switzerland.

### RSV-A F gene amino acid substitutions identified in wastewater samples of the 2023-2024 season

Mutations identified in the RSV-A F gene sequences from wastewater corresponded with the signature mutations of A.D.1, A.D.2, A.D.3, and A.D.5, and their sub-lineages (Figure 5 C,D). Signature amino acid substitutions were identified in multiple wastewater samples from Zurich and Geneva, including T12I as signature for A.D.3 and A.D.3.1, L20F as signature for A.D.3.1, and T122A as signature for A.D.5.1 and A.D.5.2. Additionally, non-signature low-frequency mutations L20I, A23T, T91R, A103T, M115T, S377N, and A518V were identified in up to five wastewater samples (Figure 5 C,D). Among these, T91R, A103T, M115T, S377N, and A518V were also detected in recent Swiss clinical samples. With the exception of T91R, all were found in global clinical sequences (Table S6).

## Discussion

Every year, severe RSV infections place a substantial burden on healthcare, primarily due to hospitalizations of infants and the elderly. The recent approval of new vaccines and monoclonal antibodies offers hope to alleviate this burden. However, their introduction also results in a change in selective pressure on the virus, highlighting the importance of RSV genomic surveillance to monitor mutations that may cause immune escape and track circulating lineages. Despite this need, clinical RSV surveillance remains insufficient in many countries, limiting our understanding of its molecular epidemiology. Wastewater-based surveillance presents a complementary approach to fill the current gaps in RSV surveillance.

Identification of RSV lineages has typically focused on the attachment glycoprotein (G) gene, which exhibits the highest genetic variability compared to other genes in the RSV genome^22,47,48^. A more comprehensive understanding of RSV evolution has been achieved by incorporating the fusion glycoprotein (F) gene - the primary target of monoclonal antibodies and vaccines against RSV - or leveraging the complete RSV genome^41,47,49,50^. In this study, we demonstrated that wastewater can serve as a valuable resource for RSV surveillance, providing population-wide insights into circulating RSV strains that mirror the diversity of clinical RSV strains. Additionally, we demonstrate that near-complete genome coverage for RSV-A and RSV-B can be obtained from wastewater, enabling the identification of signature mutations associated with lineages and deconvolution into relative abundances of RSV lineages.

The two antigenic RSV subtypes, RSV-A and RSV-B, typically alternate dominance every one or two years^22,23^. Indeed, in Swiss wastewater samples, we predominantly found RSV-B in the 2022-2023 season, followed by RSV-A in the 2023-2024 season. Multiple clinical studies have shown that the unprecedented RSV epidemic of 2022-2023, which happened after the COVID-19 pandemic, was driven by RSV-B^51–53^. The RSV-B lineage B.D.4.1.1 predominantly circulated before the COVID-19 pandemic, whereas after the pandemic, its descendant B.D.E.1 became predominant in co-circulation with B.D.4.1.1^53–56^. This is consistent with our observations in wastewater samples collected during the 2022-2023 season, with B.D.E.1 being the predominant strain and B.D.4.1.1 being present only in a very small fraction. Compared to RSV-B in the 2022-2023 season, the wastewater samples collected during the 2023-2024 season exhibited greater genomic diversity in RSV-A, with observed mutations belonging to multiple RSV-A lineages. The lineage deconvolution showed the co-circulation of A.D.1, A.D.3, and A.D.5-derived lineages. This is also consistent with clinical data, showing that multiple RSV-A lineages were co-circulating during the 2023-2024 season, with A.D.1, A.D.3, and A.D.5-derived lineages being the most prevalent in recent clinical samples^52,53,56,57^.

This study further demonstrates the potential to detect mutations in the RSV F gene from wastewater samples, an important finding given that many RSV immunoprophylaxis strategies target the F protein. Resistance-mediating mutations in the F gene can affect the effectiveness of these interventions. Notably, multiple amino acid substitutions identified in wastewater-derived F protein sequences were defined as signatures for circulating RSV-A and RSV-B lineages. For RSV-B, the S211N substitution, a signature of B.D.E.1, is located within the antigenic site targeted by Nirsevimab. However, S211N does not reduce the antibody’s neutralization potency^58–60^. Additionally, substitution R209Q, not a signature of B.D.E.1 but of multiple other lineages, including B.D, B.D.1, B.D.1.1, B.D.3, B.D.4, B.D.4.1, is also located on the antigenic site targeted by Nirsevimab, although its impact remains to be determined^61^. Two low-frequency substitutions, F12I and R42K, were also identified in wastewater. Both substitutions were also detected in clinical RSV-B sequences from the 2023-2024 season, though neither had been associated with resistance to Nirsevimab^62^. While R42K was absent from publicly available RSV-B sequences from Switzerland, it is currently present in 9.0% of all global RSV-B sequences and 34.2% of sequences assigned to the B.D.E.1 lineage on GenSpectrum^39^ (accessed 19-02-2025, numbers obtained by querying the LAPIS API^40^). Given that R42K is located on antigenic site I of the F protein^63^, it holds clinical relevance for new immunoprophylaxis targeting this site.

For RSV-A, low-frequency mutations L20I, L20F, A23T, T91R, A103T, M115T, S377N and A518V detected in wastewater have already been reported in clinical sequences^39,62^. S377N, identified in both wastewater and clinical sequences from Switzerland, resides within antigenic site III, targeted by the RSVPreF3 (AREXVY) vaccine^58^. This substitution is also present at low frequencies across multiple RSV-A lineages (A.D, A.D.1, A.D.1.5, A.D.2, A.D.3, A.D.5.2), comprising 2.9% of all sequences available on GenSpectrum (accessed 19-02-2025, numbers obtained by querying the LAPIS API). Three low-frequency non-synonymous mutations, namely RSV-B I431L and T555A, and RSV-A T91R, were detected in wastewater samples but found in either a single or no clinical sequence. This highlights the capacity of wastewater sequencing to identify locally circulating sub-lineages carrying mutations that may affect F protein antigenic sites, which might be missed by clinical genomic surveillance. As novel RSV vaccines and monoclonal antibodies targeting the F protein are rolled out in many countries during the 2024-2025 season, the 2022-2024 data from this study can serve as a baseline prior to implementation of these vaccines, offering insights into potential shifts in RSV genetic diversity driven by immunological pressure. Compared to clinical monitoring, regular wastewater-based RSV sequencing offers a more cost-effective approach to tracking circulating mutations.

Our study has several limitations. While our sampling strategy guarantees the privacy of individuals from the catchment population, it does not allow for tracing the source of RSV genetic material. As a result, it is not possible to link RSV lineages and mutations with demographic variables such as age, or epidemiological data such as vaccination status or severity of disease. Second, the relatively short size of the amplicons in our protocol have the advantage of providing better coverage in degraded samples. However, this comes with the limitation that we cannot link individual reads to reconstruct the genome of a whole virion. Third, genome coverage achieved through our amplicon-based sequencing method improves as the RSV concentration in the wastewater extract increases. Low RSV concentrations in the extract will reduce genome coverage, highlighting the importance of an efficient wastewater processing method that recovers sufficient viral material, especially during low-prevalence periods. Fourth, the mutation detection limit (frequency ≥ 0.02 in at least two wastewater samples) minimized the impact of sequencing errors but may have limited the identification of low-frequency mutations. Improved sequencing fidelity could enhance sensitivity for rare mutations. Last, wastewater-based RSV surveillance can suffer from the lack of clinical sequences that provide a reference database for deconvolution into circulating lineages. Although we used all RSV-A and RSV-B clinical sequences available on GenSpectrum to define the lineages, for RSV-A we deconvolved the signature mutation frequencies in wastewater into the RSV-A lineages that had been detected in Swiss clinical samples. However, the limited number of clinical sequences might not capture the full diversity of lineages, particularly those present at low abundances. As a result, low-abundance lineages might have been missed in the deconvolution of lineages in wastewater. Although there have been attempts to detect SARS-CoV-2 genetic variants from wastewater de novo^64,65^, comprehensive clinical genomic surveillance remains important for validating the clinical relevance of the lineages identified in wastewater.

In conclusion, we demonstrated that near-complete genome coverage of RSV-A and RSV-B can be achieved from wastewater, enabling monitoring relative abundances of lineages. While a single RSV-B lineage dominated in the 2022-2023 season, multiple RSV-A lineages co-circulated in the following season, aligning with global RSV epidemiology. Wastewater sequencing also allowed monitoring of antigenic sites targeted by vaccines, detecting both known mutations and novel low-frequency mutations that were not observed in clinical sequences from the same season. This highlights its potential to identify emerging or locally circulating variants carrying mutations that may affect F protein antigenic sites. Regular wastewater sequencing enables high-resolution monitoring of clinically relevant mutations at a fraction of the cost of equivalent clinical surveillance. Overall, wastewater-based RSV sequencing can support our understanding of RSV molecular epidemiology in large populations, providing insights into circulating strains during the introduction of novel RSV vaccines.

## Data availability

Raw sequence reads generated for this study were dehumanized based on the human reference genome (GRCh38) and submitted to ENA (in the CRAM data files format) under project number PRJEB85787, and clinical sequences used in this study under project number PRJEB83635. The bioinformatics pipeline with configuration file, primer BED files, and all scripts used for data visualization are available on Github under https://github.com/cbg-ethz/RSV-wastewater-V-pipe.

## Acknowledgements

We would like to thank members of the Wastewater Monitoring Laboratory for collecting and processing wastewater samples. We also thank Adam Lauring for sharing the RSV amplification primer sequences, as well as Isabella Eckerle for providing RSV-positive clinical samples, which we used for method optimization.

## Funding

This study was funded by the Swiss National Science Foundation (Sinergia grant CRSII5_205933). Funding for sample collection and processing was provided by the Swiss Federal Office of Public Health.

## Authors’ contributions

JdKE and AR (Methodology, Software, Validation, Formal analysis, Investigation, Data Curation, Visualization and Writing – original draft). IT (Software), IT, DD and LF (Methodology). CB (Resources and Visualization), LdP (Resources and Project administration), WF and EB (Resources and Methodology). TS, NB, TRJ (Conceptualization, Supervision and Funding acquisition). All authors (Writing – review & editing).

## Conflicts of interest

None

## Notes

### Competing Interest Statement

The authors have declared no competing interest.

### Author Declarations

The study used clinical RSV sequences, publicly available in the European Nucleotide Archive under project PRJEB83635.

