## Supplementary material for "Wastewater-based sequencing of Respiratory Syncytial Virus enables tracking of lineages and identifying mutations at antigenic sites"

**TABLES**

**Table S1. Primers and probes sequences for the RSV subtyping digital PCR assay**

| Primer name | Primer sequence |
|---|---|
| HRSV-Forward | ATGGCTCTTAGCAAAGTCAAGT |
| HRSV-Reverse | TGCACATCATAATTRGGAGTRTCA |
| HRSV-A-Probe | FAM-ATACCATCCAACGGAGCACAGGAGACAGC-BHQ1 |
| HRSV-B-Probe | HEX-ACATTAAATAAGGATCAGCTGCTGTCATCCAGCA-BHQ1 |

**Table S2. Mastermix composition and PCR parameters for the RSV subtyping digital PCR assay**

| Mastermix component | Concentration in mastermix |
|---|---|
| qScript XLT One-Step RT-qPCR ToughMix (2x) | 1X |
| HRSV-Forward | 0.5µM |
| HRSV-Reverse | 0.5µM |
| HRSV-A-Probe | 0.2µM |
| HRSV-B-Probe | 0.2µM |
| Fluorescein | 0.05µM |
| **dPCR thermocycling program** | |
| Partitioning<br>Reverse Transcriptase<br>Enzyme inactivation<br><br>45 cycles | 12min / 40°C<br>60 min / 50°C<br>10min / 95°C<br><br>30sec / 94°C 60sec / 55°C |

**Table S3. Parameters for reverse transcriptase of RSV RNA**

| **Mastermix component** | **Volume per sample** | |
|---|---|---|
| RT-LunaScript Supermix | 2µL | |
| Extract/sample | 8µL | |
| **Thermocycling conditions** | 2 minutes<br>10 minutes - 1 hour<br>1 minute | 25°C<br>55°C<br>95°C |

**Table S4. Parameters for the full-genome RSV amplification**

| **Mastermix component** | **Volume per sample** | |
|---|---|---|
| Q5 Hot Start High-Fidelity 2X Mastermix | 6.25µL | |
| Primers pool 1 or pool2 | 1.75µL | |
| cDNA | 4.5µL | |
| **Thermocycling conditions**<br><br>**35 cycles** | 30 seconds<br><br>15 seconds<br>5 minutes | 98°C<br><br>95°C<br>63°C |

**Table S5. RSV subtype -specific primer sequences for full-genome amplification**

| Primer name | Primer sequence | Fraction in mix |
|---|---|---|
| **RSV-A pool 1** | | |
| RSVA_1_LEFT | TGACAGAAGATAAAAATGGGGCAAA | 0.02 |
| RSVA_1_RIGHT | TGTATATTATGGGTGTGTGCTTGGT | 0.02 |
| RSVA_3_LEFT | ATCCAACGGAGCACAGGAGA | 0.02 |
| RSVA_3_RIGHT | CCACAATCAGGAGAGTCATGCC | 0.02 |
| RSVA_5_LEFT | GATGTTTTTGTTCATTTTGGTATAGCACAA | 0.02 |
| RSVA_5_RIGHT | AGTTGTTCAGCATATGCCTTTGC | 0.02 |
| RSVA_7_LEFT | ACATCACCCAAAGATCCCAAGA | 0.02 |
| RSVA_7_RIGHT | GGGTCCTGCACTTGCTACTACT | 0.02 |
| RSVA_9_LEFT | TCAGATGAAGTGTCTCTCAATCCA | 0.02 |
| RSVA_9_RIGHT | TGGATGATTGGAACATGGGCAC | 0.02 |
| RSVA_11_LEFT | TGCAGTCTAACATGCCTAAAATCAA | 0.02 |
| RSVA_11_RIGHT | TTGCAAATCGTGTAGCTGTGTG | 0.02 |
| RSVA_13_LEFT | CACTCCACAATCATAATCACCAACC | 0.02 |
| RSVA_13_RIGHT | TTGACTCGAGCTCTTGGTAGC | 0.02 |
| RSVA_15_LEFT | TGGGACACTCTCAATCATCTATTATTCA | 0.02 |
| RSVA_15_RIGHT | GTTGTTTTGTGGTGGGTTTGCT | 0.02 |
| RSVA_17_LEFT | AAGTGTTCAATTTTGTACCCTGCA | 0.02 |
| RSVA_17_RIGHT | GATAGCCTTCGGAGGTGGTTGA | 0.02 |
| RSVA_19_LEFT | TTACCACAATCCTTGCTGCAGT | 0.02 |
| RSVA_19_RIGHT | TCCAACACCTAACAAAAATCCAAGAA | 0.02 |
| RSVA_21_LEFT | AGTTCCAACAAAAGAACAACAGACT | 0.02 |
| RSVA_21_RIGHT | GTTTCAGCTTGTGGGAAAAAGGA | 0.02 |
| RSVA_23_LEFT | AATCGTGGGATCATAAAGACATTCTC | 0.02 |
| RSVA_23_RIGHT | TTACTTAATGTGACTGGTGTGCTTC | 0.02 |

| | | |
|---|---|---|
| RSVA_25_LEFT | GTCACGAAGGAATCCCTGCAAA | 0.02 |
| RSVA_25_RIGHT | TCGATATATGATATGACAGTATTGTACACTC | 0.02 |
| RSVA_27_LEFT | AACCCAAAAGAATCAACTGTTAGTGA | 0.02 |
| RSVA_27_RIGHT | ACATTAGCAGAATTTCCACTAATAATGGG | 0.02 |
| RSVA_29_LEFT | AGTAGACARAATCCAYTAATAGAACACAT | 0.02 |
| RSVA_29_RIGHT | TGATTGTATCTTTTTGTTTTGWGGATTG | 0.02 |
| RSVA_31_LEFT | TGGTTGTATAGTTTATCATAAGGAACTCAA | 0.02 |
| RSVA_31_RIGHT | TCTAATAATGTATGACATACTCTTGATAGCAG | 0.02 |
| RSVA_33_LEFT | AAGACAAGCMATGGATGCTGTT | 0.02 |
| RSVA_33_RIGHT | GTTCTATATAATTTTGTATGTGTGACGGC | 0.02 |
| RSVA_35_LEFT | TGTTCAGACAAGTTCAAATATTAGCAGA | 0.02 |
| RSVA_35_RIGHT | TCTATATAGTCCACTTTGCTCATCTACA | 0.02 |
| RSVA_37_LEFT | AGAGTATGCAGGWATAGGCCAC | 0.02 |
| RSVA_37_RIGHT | ACATRGGCAAATTCATATACAATGTTAATG | 0.02 |
| RSVA_39_LEFT | GATAAACTTCAAGATCTGTCAGATGATAGA | 0.02 |
| RSVA_39_RIGHT | AGTCTATGGCAGAAGTCTTTTCCA | 0.02 |
| RSVA_41_LEFT | CAATGGACATAAAATATACAACAAGCACT | 0.02 |
| RSVA_41_RIGHT | GGCTAGTATCAAAGTGATAATTTGTAGTTCT | 0.02 |
| RSVA_43_LEFT | TGTCCTAACAGAATTATTCTCATACCCAA | 0.02 |
| RSVA_43_RIGHT | ACACAAGAGATATGTCTTATAAGCATTGA | 0.02 |
| RSVA_45_LEFT | CTTGTTAGAATGGGATTGATAAATRTAGATAG | 0.02 |
| RSVA_45_RIGHT | CCTGAATGATCTATAATTTTATCAATCACAACC | 0.02 |
| RSVA_47_LEFT | TCAATTTTGTATTTAGTTCYACAGGTTG | 0.02 |
| RSVA_47_RIGHT | TGCTCCACTCTATTATAATCTTACTCCA | 0.02 |
| RSVA_49_LEFT | CCTGCAAATGTGTTCCCWGTATT | 0.02 |
| RSVA_49_RIGHT | AAGTTCATTGGTTGTCAAGCTGT | 0.02 |

| | | |
|---|---|---|
| **RSV-A pool 2** | | |
| RSVA_2_LEFT | ACAAAATATGGCACTTTCCCTATGC | 0.02 |
| RSVA_2_RIGHT | CTGTGATTAATAACATGCCRCATAAYTTAT | 0.02 |
| RSVA_4_LEFT | ACATTAGCAAGCTTAACAACTGAAATTC | 0.02 |
| RSVA_4_RIGHT | AGGCATTCATAAACAATCCTGCAA | 0.02 |
| RSVA_6_LEFT | TGGCCTAGGCATAATGGGAGAA | 0.02 |
| RSVA_6_RIGHT | TTGATGTTATAGGGCTTTCTTTGGTT | 0.02 |
| RSVA_8_LEFT | ACCACAGAAACATTTGATAACAATGAAGA | 0.02 |
| RSVA_8_RIGHT | GAAATCTTCAAGTGATAGATCATTGTCAC | 0.02 |
| RSVA_10_LEFT | TCCACATACACAGCTGCTGTTC | 0.02 |
| RSVA_10_RIGHT | TCACATAAAGCAATGATGTCATGTGT | 0.02 |
| RSVA_12_LEFT | AAGTCAATTCATAGTAGATCTTGGAGC | 0.02 |
| RSVA_12_RIGHT | AGAATTCTATRGTTATGGATGTRTTTTCCA | 0.02 |
| RSVA_14_LEFT | GCAATACTAAACAAACTCTGCGAATATAA | 0.02 |
| RSVA_14_RIGHT | TGCCAAAATAGATAATGTGATTTGTGC | 0.02 |
| RSVA_16_LEFT | ACTACATCACAATCCACCACCA | 0.02 |
| RSVA_16_RIGHT | GTGGTGTTGATGGTTGGCTTTC | 0.02 |
| RSVA_18_LEFT | AGGAAACCCTCCACTCAACCA | 0.02 |
| RSVA_18_RIGHT | ACCAACCAGTTCTTAGAGCACTAA | 0.02 |
| RSVA_20_LEFT | AGATTTATGAATTATACACTCAACAATGCC | 0.02 |
| RSVA_20_RIGHT | ACATATAAGTGCTTACAGGTGTAGTTACA | 0.02 |
| RSVA_22_LEFT | AGGATCCAACATCTGCTTAACAAGA | 0.02 |
| RSVA_22_RIGHT | CCTTCTTGCTTRTTTACATAATATAATGT | 0.02 |
| RSVA_24_LEFT | TATACTGCAAGGCCAGAAGCAC | 0.02 |
| RSVA_24_RIGHT | GCATGGGGTGGCCATTCAAA | 0.02 |
| RSVA_26_LEFT | ACTGAACTCAACAGCGATGACA | 0.02 |
| RSVA_26_RIGHT | ATTGAATGGTTGATCCGGTGGG | 0.02 |

| | | |
|---|---|---|
| RSVA_28_LEFT | CTCAAAATTTTCTACAACATMTAGGTATTACTG | 0.02 |
| RSVA_28_RIGHT | GTCATAAGTAATGAYTGAAARTAAGTAGGTTC | 0.02 |
| RSVA_30_LEFT | TGAAGACAACTCAGTCATTACTACCA | 0.02 |
| RSVA_30_RIGHT | ACATTCAATCTACTAAGGCTAATATCTTTCC | 0.02 |
| RSVA_32_LEFT | TCAGAAAACGRTTTTATAATAGTATGCTCA | 0.02 |
| RSVA_32_RIGHT | GCATTTCTTAAAGTAGGCCATCTGT | 0.02 |
| RSVA_34_LEFT | GATCTTGAAATGATCATAAATGATAARGCT | 0.02 |
| RSVA_34_RIGHT | TCTGTAGTTCTAGATCACCATATCTTGT | 0.02 |
| RSVA_36_LEFT | TCATGTCACAATAATATGCACATATAGGC | 0.02 |
| RSVA_36_RIGHT | CCGTTATGTTGGATCGTTTTACTCA | 0.02 |
| RSVA_38_LEFT | ATATAGAGGTGAAAGTCTATTATGCAGTTT | 0.02 |
| RSVA_38_RIGHT | ACGAATTCAGCATTGGGGTTTT | 0.02 |
| RSVA_40_LEFT | TGAAAGTTTACCYTTTTATAAAGCAGAGA | 0.02 |
| RSVA_40_RIGHT | CCCAKGGTTTAGTGGGTCCTCT | 0.02 |
| RSVA_42_LEFT | AGGATGAATTTATGGAGGAACTTAGCA | 0.02 |
| RSVA_42_RIGHT | CACTTGTTTTAACTTGTGAATATCAACATC | 0.02 |
| RSVA_44_LEFT | TCTCCTATGTGTATTGGAATTAATAGACAGT | 0.02 |
| RSVA_44_RIGHT | ACTTTCTAATTCRGAATTAGCAATCCT | 0.02 |
| RSVA_46_LEFT | GCTTATTAAATCGTCTACAATGATTAGAACC | 0.02 |
| RSVA_46_RIGHT | TGATCATTGCAATCTTTCAGACTTCTG | 0.02 |
| RSVA_48_LEFT | ATTTACATATAAAGTTTGCTGAACCTATCAGTC | 0.02 |
| RSVA_48_RIGHT | AGGGTATCAAACTCTTAATATTTGCATCA | 0.02 |
| RSVA_50_LEFT | GGACGTAATGAAGTTTTCAGCA | 0.02 |
| RSVA_50_RIGHT | AAACCAATTAGATTTGGATTTAAACTT | 0.02 |

| Primer name | Primer sequence | Fraction in mix |
|---|---|---|
| **RSV-B pool 1** | | |
| RSVB_1_LEFT | TCAGAAATGGGGTGCAATTCACT | 0.006 |
| RSVB_1_RIGHT | TGAATGGAGATCAAGCCCAAGT | 0.006 |
| RSVB_3_LEFT | CTCACCAAAGAAATCATCACACACA | 0.025 |
| RSVB_3_RIGHT | GCCATCTTTGTATTTGCCCCAA | 0.025 |
| RSVB_5_LEFT | GGTATGCTATTAATCACTGAAGATGCA | 0.007 |
| RSVB_5_RIGHT | TTTTTAAGACATTGTTTGCCCTCCT | 0.007 |
| RSVB_7_LEFT | TCATGCTAGGACATGCTAGTGTC | 0.015 |
| RSVB_7_RIGHT | TCAGGTGCAAACTTCTCCATGT | 0.015 |
| RSVB_9_LEFT | ATCAATCCAACAAGTGAAGCCG | 0.003 |
| RSVB_9_RIGHT | TCTTGCCATGGCCTCTAACCTA | 0.003 |
| RSVB_11_LEFT | ACGATAGCGACAATGATCTATCACT | 0.012 |
| RSVB_11_RIGHT | GTCCTTTGGGCGTAGAGATCTG | 0.012 |
| RSVB_13_LEFT | ACAGTCAAAGATCTAACCATGAAGACA | 0.016 |
| RSVB_13_RIGHT | ATGTGAACCTGTCATTCAATGTTGA | 0.016 |
| RSVB_15_LEFT | TCTAAACATCATAAACATCTACACTACACA | 0.012 |
| RSVB_15_RIGHT | GCATAATGGGAGAACACTATGTGTC | 0.012 |
| RSVB_17_LEFT | AATCTATAGCACAAATAGCACTATCAGTT | 0.005 |
| RSVB_17_RIGHT | TGCCACATATACTACAGGGAACAA | 0.005 |
| RSVB_19_LEFT | CACAATCCACTGTGCTCGACA | 0.036 |
| RSVB_19_RIGHT | ACCTCTGCTAACTGCACTACATG | 0.036 |
| RSVB_21_LEFT | ACAGTTACTTATGCAAAACACACCAG | 0.008 |
| RSVB_21_RIGHT | AATCTGCTGTTCTTCTGCTGGA | 0.008 |
| RSVB_23_LEFT | TCAATGATATGCCTATAACAAATGATCAGA | 0.008 |
| RSVB_23_RIGHT | ACTGAGCTGCTTATGTCTGTTTTTG | 0.008 |
| RSVB_25_LEFT | GACATTTTCTAATGGCTGTGATTATGTG | 0.008 |

| | | |
|---|---|---|
| RSVB_25_RIGHT | GCTGAATGCAATATTATTGATTCCACTT | 0.008 |
| RSVB_27_LEFT | GCGAAGAAATCCCTGCAAATTTG | 0.006 |
| RSVB_27_RIGHT | AGATGGATGGTTTGCTTGCTGT | 0.006 |
| RSVB_29_LEFT | CCATAAGTGATCAAAATGACCAAACC | 0.036 |
| RSVB_29_RIGHT | GAGATAACACCTTTTAGATAACTATCAGTTAGA | 0.036 |
| RSVB_31_LEFT | ACACAGTCATTAATATCTAGATACCATAAAGG | 0.023 |
| RSVB_31_RIGHT | TGAACCAGTGTATTAACCATGATGGA | 0.023 |
| RSVB_33_LEFT | GAAAGACATCAGCCTCAGYAGA | 0.036 |
| RSVB_33_RIGHT | TTGAGATTATTATCACCTGCAAGCTT | 0.036 |
| RSVB_35_LEFT | CCTACCTCTAAGATGGTTAAACTATTATAAACT | 0.022 |
| RSVB_35_RIGHT | AGCAAACATTCTACCCACACTGA | 0.022 |
| RSVB_37_LEFT | TCAAGCATTTAGATATGAAACATCATGTGT | 0.036 |
| RSVB_37_RIGHT | GCATACTCTTTATATAGCAACTTAAGGCT | 0.036 |
| RSVB_39_LEFT | CAGAGGAGAAAGCTTATTATGCAGTTT | 0.017 |
| RSVB_39_RIGHT | AATGTTACAAACTCGGCATTTGGA | 0.017 |
| RSVB_41_LEFT | GTGCGCAACACTATACTACCACT | 0.019 |
| RSVB_41_RIGHT | AGTGCTAGTTGTATATTTAATGTCCATTGT | 0.019 |
| RSVB_43_LEFT | ACTGAGTACTGGAACACTTGGAC | 0.023 |
| RSVB_43_RIGHT | TTGGGTTAAACTTATTTTATCTGGTAGGA | 0.023 |
| RSVB_45_LEFT | TGTTTTCATAGAGGTTATGGTAAAGCA | 0.025 |
| RSVB_45_RIGHT | CAGATGAGTGTTGTCTGAAAAGTTGT | 0.025 |
| RSVB_47_LEFT | CTAACAAAACAAATAAGGATTGCYAATTC | 0.036 |
| RSVB_47_RIGHT | TGATCTTGCATCCTGTGGAACT | 0.036 |
| RSVB_49_LEFT | ACCATTCCTGCTACAGATGCAA | 0.024 |
| RSVB_49_RIGHT | TTGCATCRATAGATTCCTTGTCAATCT | 0.024 |
| RSVB_51_LEFT | CAGYAACAAGCTTATAAACCACA | 0.036 |
| RSVB_51_RIGHT | AAATGCACATGTGTGATTGTTAG | 0.036 |

| | | |
|---|---|---|
| **RSV-B pool 2** | | |
| RSVB_2_LEFT | ACACACTGCTCTCAATTAAATGGTC | 0.006 |
| RSVB_2_RIGHT | TGACTAGGAATGTAAATGTAGCTTGTCT | 0.006 |
| RSVB_4_LEFT | TCCTATAATATACAAATATGACCTCAACCC | 0.022 |
| RSVB_4_RIGHT | AAGTATCTTTATAGTGTCTTCCCTTCCT | 0.022 |
| RSVB_6_LEFT | ACTGTGTATAGCTGCCCTTGTAA | 0.004 |
| RSVB_6_RIGHT | GGTAGAAACCAGCTTCTCCTCC | 0.004 |
| RSVB_8_LEFT | GCAACTCAAAGAAAATGGAGTAATAAACT | 0.008 |
| RSVB_8_RIGHT | TGGGGTGAGATCTTCTTTGAAGC | 0.008 |
| RSVB_10_LEFT | GAGGAGATCAATGACCAAACAAATGA | 0.004 |
| RSVB_10_RIGHT | TGATTCAATGGATTGGTGTCTGTTT | 0.004 |
| RSVB_12_LEFT | ATGTTCCAGTCATCTGTGCCAG | 0.005 |
| RSVB_12_RIGHT | TGCTATATTTTCCAGTGAGTTCAGATC | 0.005 |
| RSVB_14_LEFT | TGGATCTTGGTGCTTACCTAGAAA | 0.011 |
| RSVB_14_RIGHT | GTGTAAAATAGGGCCAAAATTTGCTT | 0.011 |
| RSVB_16_LEFT | GCTAAGTGAACATAAAATATTCTGCAACA | 0.023 |
| RSVB_16_RIGHT | TGGCTGCAATTATAAGAGAGGTTGA | 0.023 |
| RSVB_18_LEFT | GGCACAACCTCTACTCCAACAC | 0.004 |
| RSVB_18_RIGHT | TGTTTTGATGTGGTTGTGTCGAG | 0.004 |
| RSVB_20_LEFT | TGCATTGTACCTTACCTCAAGTCA | 0.009 |
| RSVB_20_RIGHT | TCCTTCAAGGTGTAGAACTTTGGA | 0.009 |
| RSVB_22_LEFT | TGTTTTAACCAACAGAGTGTTAGATCT | 0.011 |
| RSVB_22_RIGHT | CTTCTTTGATGTTGGTGGTGCA | 0.011 |
| RSVB_24_LEFT | AGTGAAGTCAGCCTTTGTAACACT | 0.011 |
| RSVB_24_RIGHT | ACATTATGTAATAATTCATCGGATCTACGA | 0.011 |
| RSVB_26_LEFT | AGCTATTGGTTTACTGTTGTATTGCA | 0.037 |
| RSVB_26_RIGHT | AATGCATGAGGAGGCCATTCAA | 0.037 |
| RSVB_28_LEFT | TGATAGAACAGAAGAATATGCTCTTGGT | 0.009 |
| RSVB_28_RIGHT | TGAGATCAATATGGATGATATACTACAAGGA | 0.009 |

| | | |
|---|---|---|
| RSVB_30_LEFT | CATCTTAACATCCCTGAAGATATATATACAGT | 0.037 |
| RSVB_30_RIGHT | TTTGTTCAGACGAGGACATGCT | 0.037 |
| RSVB_32_LEFT | ACTCTCAGCTGTGGAAAACAATCA | 0.037 |
| RSVB_32_RIGHT | AATCCGCATCTCAACCCTAAGC | 0.037 |
| RSVB_34_LEFT | ACAAGACAGTGTCTGATAATATCATAAATGG | 0.020 |
| RSVB_34_RIGHT | ACTCACGATAGAACCGCAATCC | 0.020 |
| RSVB_36_LEFT | TGTGTAGTTAATCAAAGCTATCTCAACA | 0.024 |
| RSVB_36_RIGHT | ACAAGAGGTATTGTTAAATGCAACCA | 0.024 |
| RSVB_38_LEFT | TGATAAATGGTGATAACCAGTCAATTGA | 0.037 |
| RSVB_38_RIGHT | GCTTATTATTACATAAAGCATGATTTCGGA | 0.037 |
| RSVB_40_LEFT | CTGTTTGGTGGTGGTGATCCTA | 0.024 |
| RSVB_40_RIGHT | CATAAACAACTCTTAATCCATGAGGGT | 0.024 |
| RSVB_42_LEFT | GCAAGTATGTAAGAGAAAGATCTTGGT | 0.037 |
| RSVB_42_RIGHT | CGCATGGTCTACTACTGACTGTT | 0.037 |
| RSVB_44_LEFT | TGATGAAGCCTCCTATATTTACAGGA | 0.037 |
| RSVB_44_RIGHT | AACACAAAGAAGATCTGAAGTGTTCA | 0.037 |
| RSVB_46_LEFT | TGCTAAATTTACCGTATGCCCTTG | 0.019 |
| RSVB_46_RIGHT | TGGTAACAGTGGAAGATTTAATATGCA | 0.019 |
| RSVB_48_LEFT | AAGGAAYAGTGCATCACTTTATTGCA | 0.037 |
| RSVB_48_RIGHT | CTCCAATTGGCTGTGACAGGTA | 0.037 |
| RSVB_50_LEFT | GGCCCTGCAAATATACTTCCTGT | 0.024 |
| RSVB_50_RIGHT | ACACTACCTGTTATTTTAATCAGCTTCT | 0.024 |

**Table S6. Prevalence of amino acid substitutions in the F gene in clinical sequences from Switzerland and globally.**

| **Substitution in RSV-A F gene** | **Percentage of Swiss sequences with mutation*** *(sequences with mutations/sequences with non-ambiguous nucleotide at that position)* | **Percentage of global sequences with mutation#** *(sequences with mutations/sequences with non-ambiguous nucleotide at that position)* |
|---|---|---|
| T12I | 19.1% (9/47) | 14.0% (1,393/9,943) |
| L20I | 0.0% | 0.1% (10/9,953) |
| L20F | 8.5% (4/47) | 2.1% (210/9,953) |
| A23T | 0% | 3.6% (361/9,946) |
| T91R | 2.2% (1/46) | 0% |
| A103T | 8.3% (4/48) | 8.7% (867/9,988) |
| M115T | 4.4% (2/46) | 0.8% (84/10,000) |
| T122A | 30.4% (14/46) | 22.3% (2230/9,991) |
| S377N | 27.3% (12/44) | 2.9% (287/9,906) |
| A518V | 6.7% (3/45) | 4.2% (415/9,925) |
| **Substitution in RSV-B F gene** | **Percentage of Swiss sequences with mutation#** | **Percentage of global sequences with mutation#** |
| F12I | 50.0% (4/8) | 2.5% (213/8,468) |
| R42K | 0.0% | 9.0% (766/8,477) |
| S190N | 87.5% (7/8) | 27.8% (2,342/8,433) |
| R209Q | 0.0% | 23.7% (2,001/8,431) |
| S211N | 87.5% (7/8) | 27.8% (2,340/8,429) |
| S389P | 87.5% (7/8) | 26.1% (2,181/8,370) |
| I431L | 0.0% | 0.0% |
| T555A | 0.0% | 0.0% (1/8,342) |

*Based on publicly available mutation annotation of the sequences, using the Nextclade tool.
#Based on publicly available sequences from GenSpectrum (accessed 19-02-2025), numbers obtained by querying the LAPIS API.

**FIGURES**

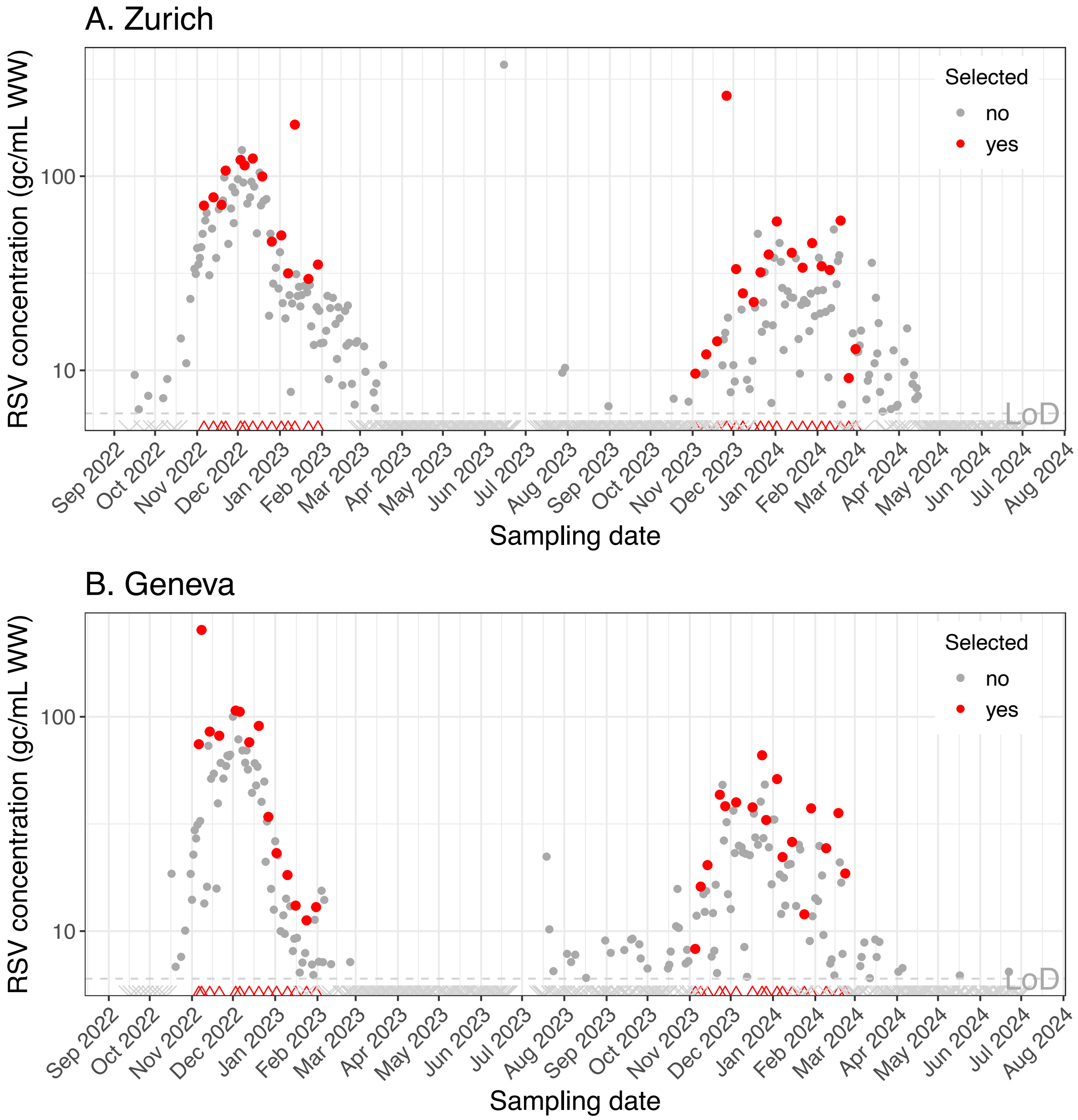


**Figure S1. Selection of wastewater samples for RSV sequencing, obtained from wastewater treatment plants in Zurich (A) or Geneva (B).** Pan-RSV concentrations have been measured since September 2022 as part of the Swiss wastewater-based respiratory virus monitoring program. During the seasonal RSV waves, one extract per location per week was selected for sequencing, prioritizing the extract with the highest or second-highest measured RSV concentration. Samples selected for sequencing are highlighted with red dots, with their collection dates additionally marked as red triangles on the x-axis. The limit of detection (LoD) for the pan-RSV digital PCR assay was six genome copies per mL of wastewater, corresponding to at least three positive partitions in the digital PCR, and is represented by a dashed grey line. Samples with pan-RSV concentrations below the LoD are shown as grey crosses on the x-axis.

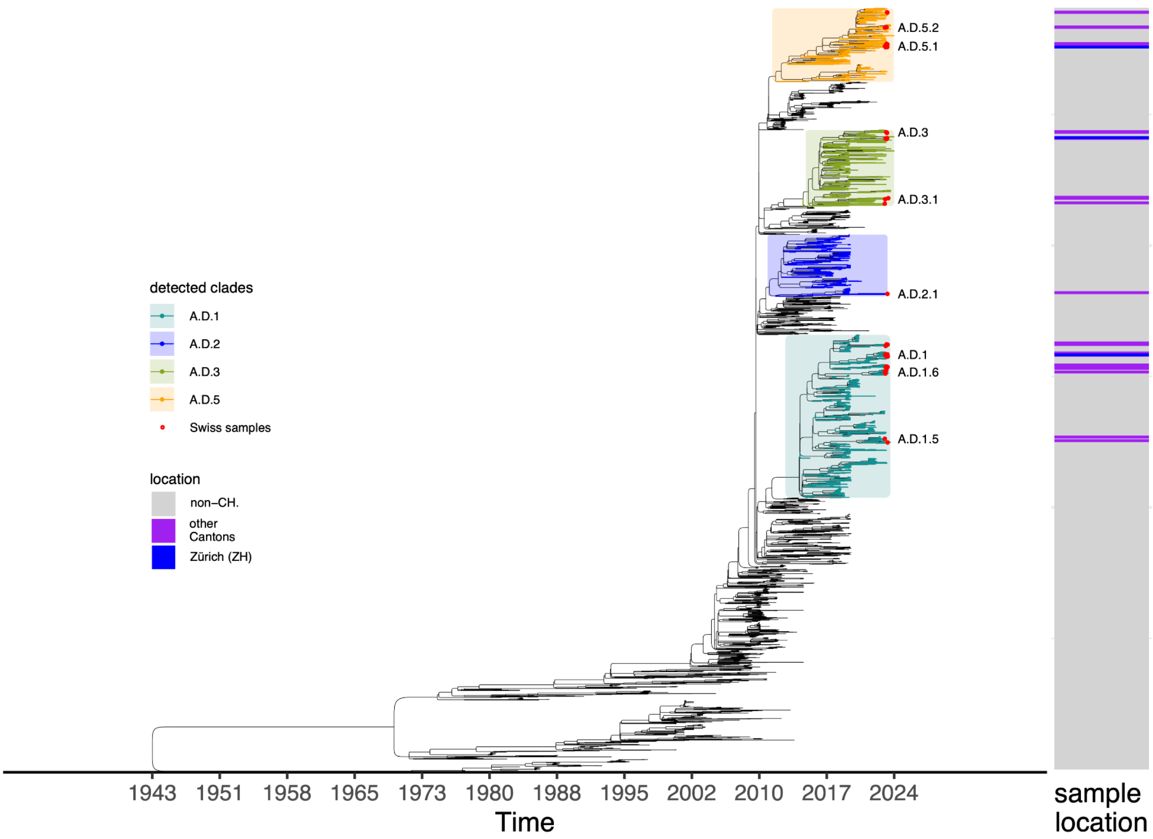


**Figure S2. Phylogenetic analysis of Swiss clinical RSV-A sequences.** Phylogenetic analysis performed with the adapted Nextstrain RSV-A build detected eight RSV-A lineages (A.D.1, A.D.1.5, A.D.1.6, A.D.2.1, A.D.3, A.D.3.1, A.D.5.1, A.D.5.2) circulating in Switzerland during the 2023-2024 season. Sequences from Switzerland (whole genome non-N coverage > 0.66, n=51) are highlighted in the phylogeny with red tips and labeled with their detected lineages. Detected clades are highlighted directly on the tree. The samples' locations are visualized on the right: samples from the canton of Zurich are highlighted in dark blue, from Swiss cantons other than Zurich or Geneva in purple, and samples originating from outside of Switzerland in grey. Note that there were no clinical samples originating from the canton of Geneva.

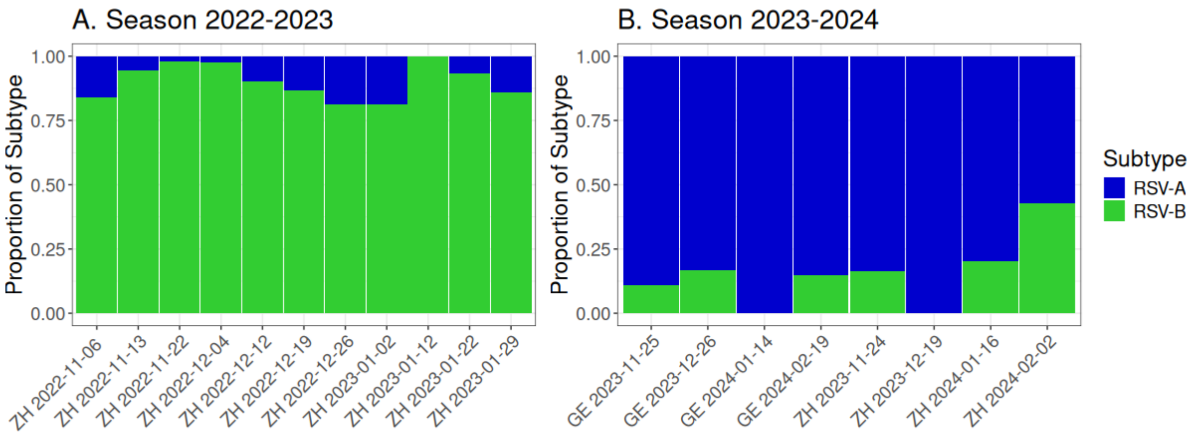


**Figure S3. Results of RSV subtyping using an RSV-A and RSV-B discriminating digital PCR assay.** (A) RSV positive wastewater extracts from the RSV season 2022-2023 showed dominance of RSV-B. Primers and probes are reported in Table S2, with the exception that in this experiment, the probe for detecting RSV-A was the same as reported by Wang et al[1]. (B) RSV-A was dominant in wastewater extracts obtained during the RSV season of 2023-2024. X-axis labels contain the wastewater sampling locations Zurich (ZH) or Geneva (GE) and the sampling dates. Primers and probes are reported in Table S2.

1. Wang L, Piedra PA, Avadhanula V, et al. Duplex real-time RT-PCR assay for detection and subgroup-specific identification of human respiratory syncytial virus. J Virol Methods. 2019;271:113676

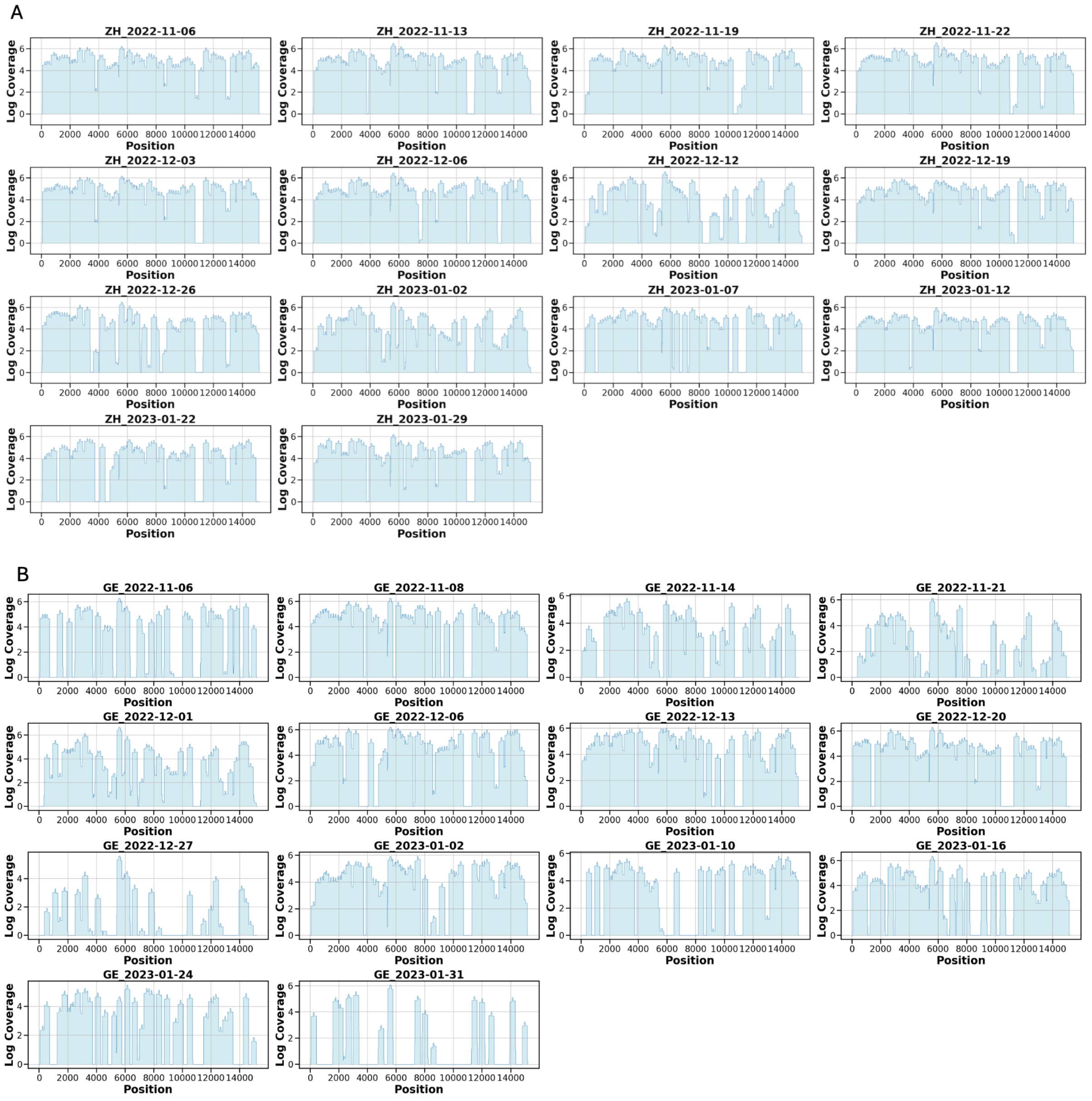


**Figure S4. RSV-B genome coverage obtained from wastewater samples from Zurich (A) Geneva (B) during the RSV season of 2022-2023.** Position of the genome is represented on the horizontal axis, genome coverage (in log scale) is provided on the vertical axis. Coverage was estimated after trimming and filtering of the reads.

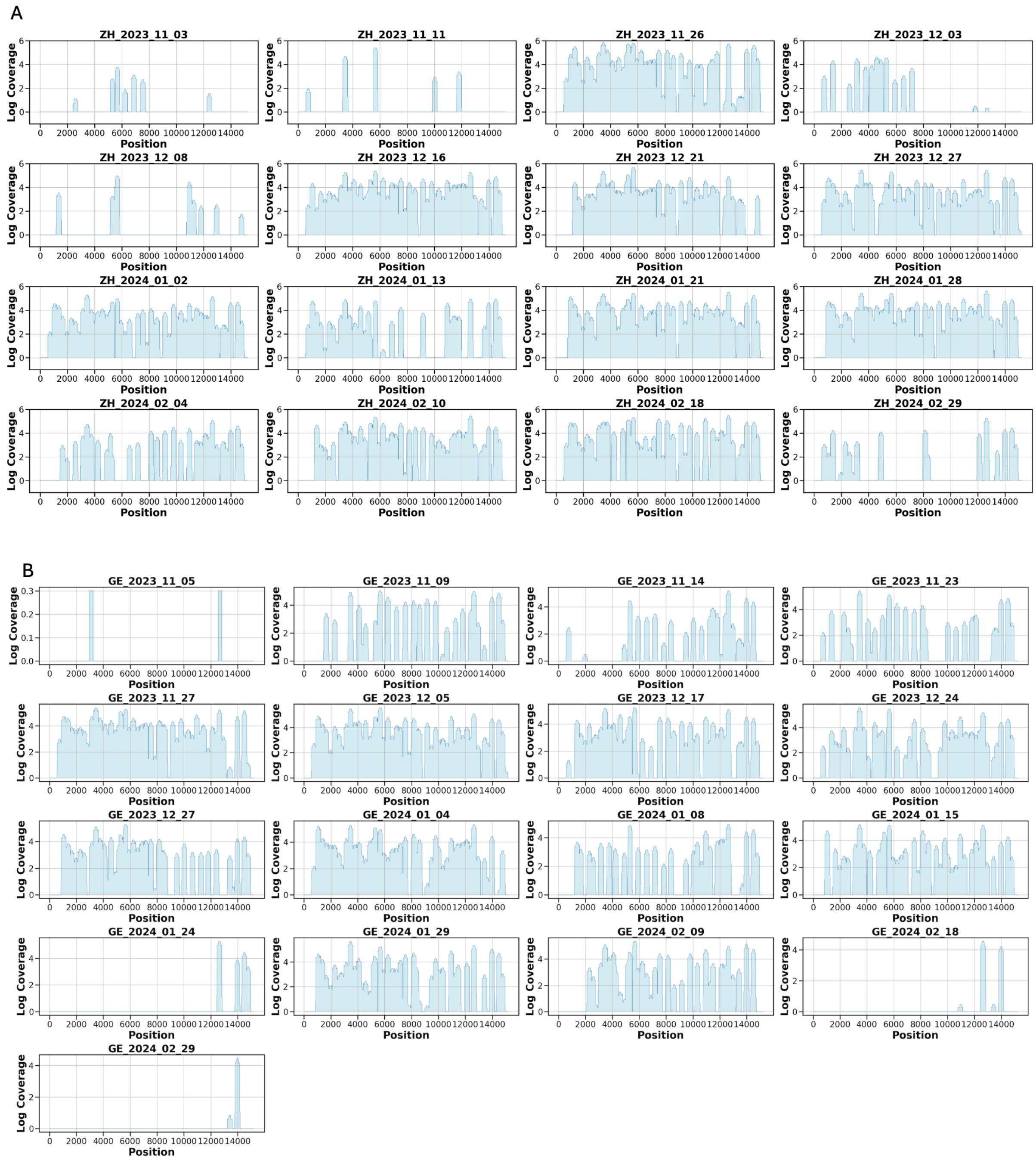


**Figure S5. RSV-A genome coverage obtained from wastewater samples from Zurich (A) Geneva (B) during the RSV season of 2023-2024.** Position of the genome is represented on the horizontal axis, genome coverage (in log scale) is provided on the vertical axis. Coverage was estimated after trimming and filtering of the reads. Two samples from Zurich on dates 2024-02-24 and 2023-11-19 and a single sample from Geneva on 2024-02-23 had no genome coverage and are not shown.

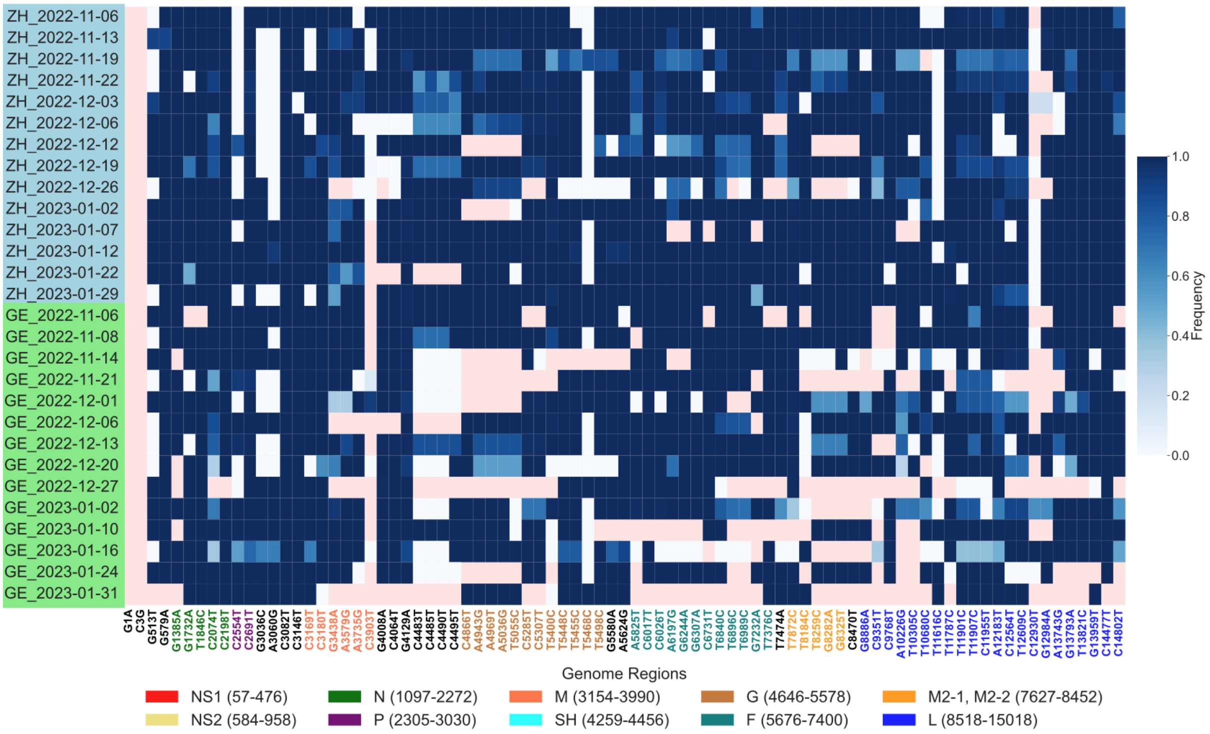


**Figure S6. Frequencies of all signature mutations of the RSV-B lineage B.D.E.1 in the wastewater samples.** Observed frequencies of all B.D.E.1 signature mutations (synonymous and non-synonymous) in wastewater-derived RSV-B sequences from Zurich (ZH, blue highlighted row labels) and Geneva (GE, green highlighted row labels) obtained during the RSV season of 2022-2023. Rows refer to samples and labels include the date and location of collection. Frequencies are encoded from white (lowest, 0.0) to dark blue (highest, 1.0), while dropouts (read depth below 30 per position) are shown in pink. Mutations are provided in terms of nucleotide substitutions relative to the reference EPI_ISL_1653999 and color-coded according to the gene in which they occur.
